# Clinical characteristics of patients hospitalized with COVID-19 in Spain: results from the SEMI-COVID-19 Network

**DOI:** 10.1101/2020.05.24.20111971

**Authors:** José Manuel Casas Rojo, Juan Miguel Antón Santos, Jesús Millán Núñez-Cortés, Carlos Lumbreras, José Manuel Ramos Rincón, Emilia Roy-Vallejo, Arturo Artero, Francisco Arnalich Fernández, Jose Miguel García Bruñén, Juan Antonio Vargas Núñez, Santiago J. Freire Castro, Luis Manzano, Isabel Perales Fraile, Anxela Crestelo Vieitez, Francesc Puchades, Enrique Rodilla, Marta Nataya Solís Marquínez, David Bonet Tur, María del Pilar Fidalgo Moreno, Eva M Fonseca Aizpuru, Francisco Javier Carrasco Sánchez, Elisa Rabadán Pejenaute, Manuel Rubio-Rivas, José David Torres Peña, Ricardo Gómez Huelgas, for the SEMI-COVID-19 Network

## Abstract

**Background:** Spain has been one of the countries most affected by the COVID-19 pandemic.

**Objective:** To create a registry of patients with COVID-19 hospitalized in Spain in order to improve our knowledge of the clinical, diagnostic, therapeutic, and prognostic aspects of this disease.

**Methods:** A multicentre retrospective cohort study, including consecutive patients hospitalized with confirmed COVID-19 throughout Spain. Epidemiological and clinical data, additional tests at admission and at seven days, treatments administered, and progress at 30 days of hospitalization were collected from electronic medical records.

**Results:** Up to April 30^th^ 2020, 6,424 patients from 109 hospitals were included. Their median age was 69.1 years (range: 18-102 years) and 56.9% were male. Prevalences of hypertension, dyslipidemia, and diabetes mellitus were 50.2%, 39.7%, and 18.7%, respectively. The most frequent symptoms were fever (86.2%) and cough (76.5%). High values of ferritin (72.4%), lactate dehydrogenase (70.2%), and D-dimer (61.5%), as well as lymphopenia (52.6%), were frequent. The most used antiviral drugs were hydroxychloroquine (85.7%) and lopinavir/ritonavir (62.4%). 31.5% developed respiratory distress. Overall mortality rate was 21.1%, with a marked increase with age (50-59 years: 4.2%, 60-69 years: 9.1%, 70-79 years: 21.4%, 80-89 years: 42.5%, ≥ 90 years: 51.1%).

**Conclusions:** The SEMI-COVID-19 Network provides data on the clinical characteristics of patients with COVID-19 hospitalized in Spain. Patients with COVID-19 hospitalized in Spain are mostly severe cases, as one in three patients developed respiratory distress and one in five patients died. These findings confirm a close relationship between advanced age and mortality.

## INTRODUCTION

Spain is one of the countries with the highest number of patients with severe acute respiratory syndrome coronavirus 2 (SARS-CoV-2) in the world. Since the first COVID-19 infection was confirmed in the country on January 31, 2020, 222,857 cases have been diagnosed and 26,299 patients have died.[1]

Current knowledge about COVID-19 is incomplete and fragmented. Cohort studies from various countries [2-7] suggest that the risk factors and prognosis of this disease may not be able to be extrapolated to other geographical areas, as they could be influenced by specific public health conditions or race-related issues. To date, there are no solid therapeutic recommendations, as the results from ongoing clinical trials on the efficacy of antiviral and immunosuppressant drugs are pending [8,9].

The SEMI-COVID-19 Network arises as an initiative of the Spanish Society of Internal Medicine (SEMI) to improve the quality of treatment for SARS-CoV-2. The main objective of the registry is to generate, in a short period of time, a large, multicenter cohort with detailed information on the epidemiology, clinical progress, and treatment received by patients. This will allow for the development of prognostic models and the assessment of the efficacy of different treatment regimens used in real-world clinical practice.

## METHODS

### Study design

Observational study.

The SEMI-COVID registry is an ongoing retrospective cohort, comprising most consecutive patients hospitalized in Spain from March 1^st^ 2020 up to the end of the pandemy, discharged with confirmed COVID-19 disease. Inclusion was started as of March 24th and is still ongoing. Follow-up at one month was done via telephone.

### Study population and participants

All consecutive patients discharged or dead after hospital admission, with confirmed SARS-COV-2 infection, were eligible for inclusion. COVID-19 was confirmed either by positive result on real-time polymerase chain reaction (RT-PCR) testing of a nasopharyngeal or sputum sample, or by positive result of serological testing and a clinically compatible presentation.

Inclusion criteria for the registry were: a) patient age≥ 18 years, b) a confirmed diagnosis of COVID-19, c) first hospital admission in a Spanish Hospital participant in the study, d) hospital discharge or death at hospital.

Exclusion criteria were subsequent admissions for the same patient, and denial or withdrawal of informed consent.

Patients were treated at their attending physician’s discretion, according to local protocols and clinical judgement. Patients included in open-label clinical trial could be included in the registry, provided all information about treatment was available. This registry, by its observational characteristics, provided no additional inconvenience to the patients included.

### Registry information

An online electronic data capture system (DCS) has been developed, which includes a database manager along with procedures for the verification of data and contrasting of information against the original medical record in order to ensure the best possible quality of data collection.

Patient identifiable data are dissociated and pseudonymized. Direct identifiers are not collected in the DCS, but rather an alphanumeric sequence of characters that includes a code for identification of the researcher and a correlative number is used. Each researcher must maintain a protected registry (patient log) that is for his/her sole use. The purpose of this protected registry is to be able to confirm data with the medical records so that additional information may be gathered, if necessary, as well as to perform quality controls. This system allows for patient privacy to be respected and ethical considerations to be met while also complying with data protection regulations.

The database platform is hosted on a secure server. All information contained in the database, the configuration of the information within the database, as well as the database itself are fully encrypted. Every client-server data transfer is encrypted through a valid TLS certificate. Daily backups are performed in order to ensure data integrity.

### Data collection

Data are collected retrospectively and include approximately 300 variables grouped under various headings: (1) inclusion criteria, (2) epidemiological data, (3) RT-PCR and serology data, (4) personal medical and medication history, (5) symptoms and physical examination findings at admission, (6) laboratory (blood gases, metabolic panel, complete blood count, coagulation) and diagnostic imaging tests, (7) additional data at seven days after admission or at admission to the intensive care unit, (8) pharmacological treatment during the hospitalization (antiviral drugs, immunomodulators, antibiotics) and ventilator support, (9) complications during the hospitalization, and (10) progress after discharge and/or 30 days from diagnosis. A list of variables can be found in the Appendix 1.

### Study Management

The Spanish Society of Internal Medicine (SEMI, for its initials in Spanish) is the sponsor of this study. The researchers that coordinate the study from each hospital are SEMI members and were asked to participate in the study on a voluntary basis without receiving remuneration.

Database monitoring is performed by the study’s scientific steering committee and an independent external agency. Logistics coordination and data analysis are also carried out by external independent agencies.

### Data analysis

Participants’ demographic, clinical, epidemiological, laboratory, and diagnostic imaging data were analyzed. Treatment received, complications, and clinical progress were also examined. Quantitative variables are expressed as median [interquartile range]. Categorical variables are expressed as absolute frequencies and percentages. Mortality is expressed as case fatality rate (CFR).

### Ethical aspects

Personal data is processed in strict compliance with Spanish Law 14/2007, of July 3, on Biomedical Research; Regulation (EU) 2016/679 of the European Parliament and of the Council of 27 April 2016 on the protection of natural persons with regard to the processing of personal data and on the free movement of such data, and repealing Directive 95/46/EC (General Data Protection Regulation); and Spanish Organic Law 3/2018, of December 5, on the Protection of Personal Data and the Guarantee of Digital Rights. In accordance with applicable regulations, the Spanish Agency of Medicines and Medical Products (AEMPS, for its initials in Spanish) has ruled that due to its nature, the study only required the approval of the Ethics Committee and not the Autonomous Community, as in other studies. The SEMI-COVID-19 Registry has been approved by the Provincial Research Ethics Committee of Malaga (Spain).

Informed consent was obtained from all the patients. When it was not possible to obtain informed consent in writing due to biosafety concerns or if the patient had already been discharged, informed consent was requested verbally and noted on the medical record.

The STROBE statement guidelines were followed in the conduct and reporting of the study.

## RESULTS

Up to April 30, 2020, 6,424 patients hospitalized in 109 hospitals throughout Spain were included in the registry (Figure 1). The epidemiological characteristics of population studied are described in Table 1. The median age was 69.1 years (range: 18-102 years) and 56.9% were male. Male gender was predominant in all age ranges except for patients ≥90 years, in which females accounted for 56.7% of the total. A high level of multicomorbidity was observed (60.2% with moderate or severe Charlson comorbidity index scores). Furthermore, 15.1% of patients had moderate or severe dependency for activities of daily living (Barthel index score <60). The most common comorbidities were hypertension (50.2%), dyslipidemia (39.7%), obesity (21.2%), and diabetes mellitus (18.7%).

**Figure 1.**
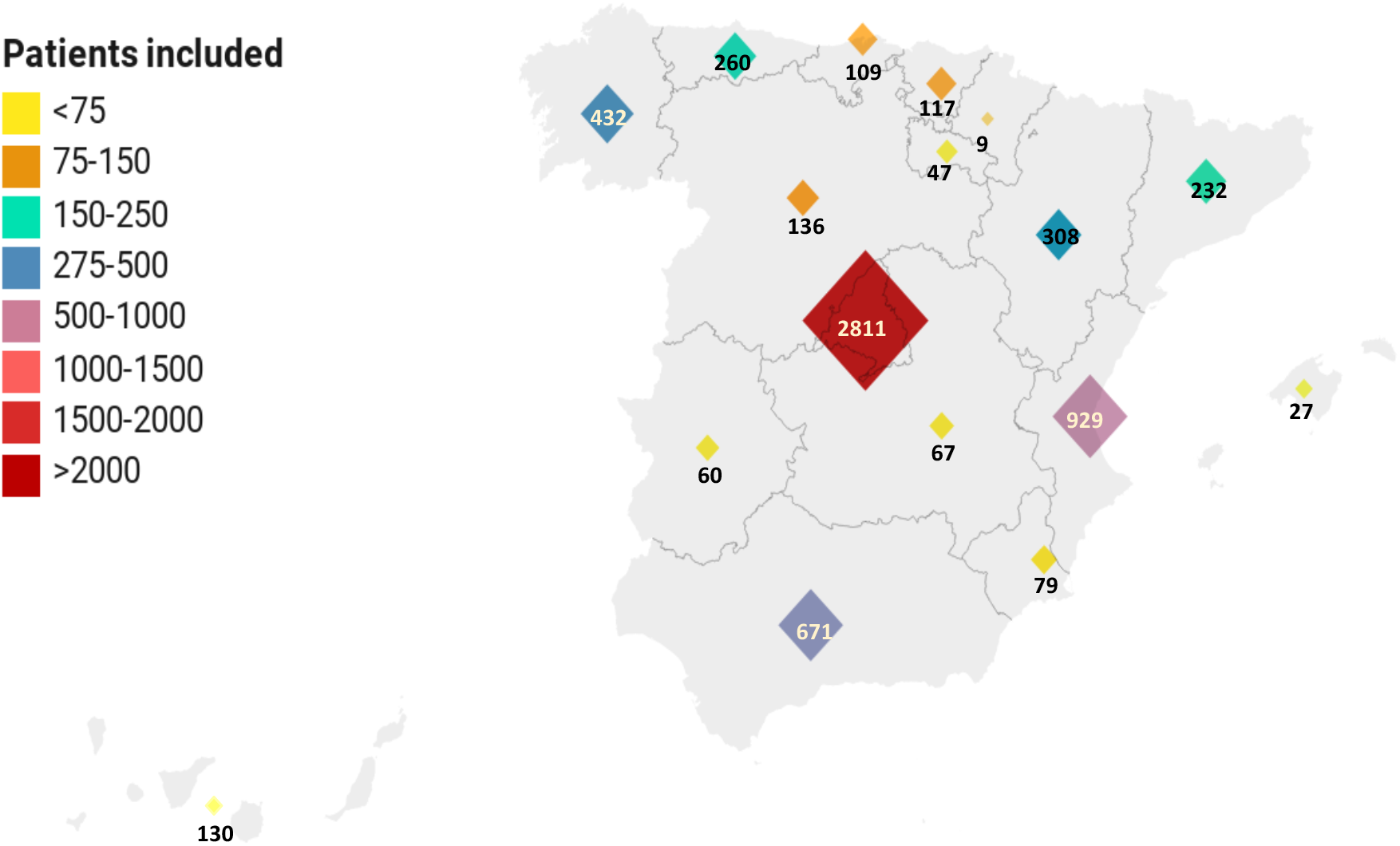
Geographical origin of patients, by Autonomous Community.

**Table 1.** Demographic and comorbidity data.

|  | Absolute frequency (%).<br>*Median [Interquartile range] | Valid cases |
| --- | --- | --- |
| Age (years) | 69.1[55.7;79.5] * | 6396 |
| 18-29 | 113 (1.8%) |  |
| 30-64 | 2583 (40.4%) |  |
| 65-79 | 2200 (34.4%) |  |
| ≥80 | 1500 (23.5%) |  |
| Gender |  | 6413 |
| Male | 3651 (56.9%) |  |
| Female | 2762 (43.1%) |  |
| Race/ethnicity |  | 6320 |
| Caucasian | 5673 (89.8%) |  |
| Other | 647 (10.2%) |  |
| Health care worker | 278 (4.3%) | 6396 |
| Age-adjusted Charlson Comorbidity Index score |  | 6107 |
| No comorbidities | 778 (12.7%) |  |
| Mild | 1655 (27.1%) |  |
| Moderate | 1649 (27%) |  |
| Severe | 2025 (33.2%) |  |
| Degree of dependency |  | 6298 |
| Independent or mild | 5285 (83.9%) |  |
| Moderate | 580 (9.2%) |  |
| Severe | 433 (6.9%) |  |
| Tobacco use |  | 6165 |
| Has never smoked | 4198 (68.1%) |  |
| Former smoker | 1643 (26.7%) |  |
| Smoker | 324 (5.3%) |  |
| Alcohol use disorder | 295 (4.7%) | 6280 |
| Obesity (BMI ≥30 kg/m<sup>2</sup>) | 1207 (21.2%) | 5691 |
| Hypertension | 3215 (50.2%) | 6400 |
| Dyslipidemia | 2540 (39.7%) | 6402 |
| Diabetes mellitus | 1193 (18.7%) | 6377 |
| Malignancy (solid tumor, leukemia, lymphoma) | 684 (10.7%) | 6366 |
| Cardiovascular disease (myocardial infarction, angina pectoris, heart failure, atrial fibrillation): | 1322 (20.7%) | 6381 |
| Angina pectoris | 243 (3.8%) | 6393 |
| Atrial fibrillation | 748 (11.7%) | 6393 |
| Heart failure | 509 (8.0%) | 6391 |
| Myocardial infarction | 426 (6.7%) | 6397 |
| Obstructive lung disease (COPD, asthma) | 992 (15.5%) | 6381 |
| COPD | 489 (7.7%) | 6390 |
| Asthma | 537 (8.4%) | 6388 |
| Obstructive sleep apnea/hypopnea syndrome | 420 (6.6%) | 6353 |
| Known HIV infection | 40 (0.6%) | 6370 |
| Moderate-severe kidney disease | 387 (6.1%) | 6385 |

Footnote: BMI: body mass index

Table 2 summarizes the clinical and radiological findings upon admission to the emergency department. The most common clinical manifestations were fever (86.2%), cough (76.5%), dyspnea (57.6%), and asthenia (47.5%). Anosmia, dysgeusia, and hyporexia were less common. Gastrointestinal manifestations were quite common, especially diarrhea. At triage, only 52.9% of patients were febrile and almost half showed some degree of respiratory impairment (oxygen saturation <90% in 16.3%, respiratory rate >20 breaths per minute in 30.2%). The qSOFA score was ≥2 in just 9.3% of patients. Lung involvement was less common upon examination than in the radiographic findings: rales were present in 52.4% of patients whereas pneumonia or interstitial infiltrates were observed on plain chest X-rays in 86.6% of patients.

**Table 2.** Clinical, laboratory, and diagnostic imaging findings at emergency room admission.

|  | Absolute frequency (%).<br>*Median [Interquartile<br>range] | Valid<br>cases |
| --- | --- | --- |
| **Clinical presentation** |  |  |
| Fever |  | 6385 |
| No | 945 (14.8%) |  |
| Mild (temperature <38 °C) | 1353 (21.2%) |  |
| Severe (temperature ≥38 °C) | 4087 (64.0%) |  |
| Cough |  | 6386 |
| No | 1501 (23.5%) |  |
| Dry | 3814 (59.7%) |  |
| Productive | 1071 (16.8%) |  |
| Fatigue | 2965 (47.5%) | 6236 |
| Diarrhea | 1418 (22.5%) | 6313 |
| Anorexia | 1390 (22.4%) | 6201 |
| Shortness of breath | 3666 (57.6%) | 6366 |
| Anosmia | 385 (6.3%) | 6109 |
| **Physical examination** |  |  |
| Oxygen saturation (pulse oximetry) (%) | 94[91;97] * | 6246 |
| <90 | 1021 (16.3%) |  |
| ≥90 | 5225 (83.7%) |  |
| O<sub>2</sub> saturation/FiO<sub>2</sub> ratio (%) | 442.9 [408.3;457.1] * | 6378 |
| Temperature (°C) | 37 [36.4;37.8] * | 6421 |
| <37 °C | 3024 (47.1%) |  |
| 37-37.9 °C | 1990 (31.0%) |  |
| ≥38 °C | 1407 (21.9%) |  |
| Hypotension (systolic blood pressure <100 mmHg) | 375 (6.1%) | 6122 |
| Tachycardia (>100 beats per minute) | 1581 (24.6%) | 6424 |
| Tachypnea (>20 breaths per minute) | 1870 (30.2%) | 6197 |
| Confusion | 734 (11.6%) | 6324 |
| Rales | 3285 (52.4%) | 6264 |
| **Chest x-ray** |  | 6355 |
| No pulmonary infiltrates | 851 (13.4%) |  |
| Unilateral pulmonary infiltrates | 1470 (23.1%) |  |
| Bilateral pulmonary infiltrates | 4034 (63.5%) |  |
| **Complete blood count** |  |  |
| White blood cell count (x10<sup>6</sup>/L) | 6100 [4650;8200] * | 6390 |
| Absolute count (x10<sup>6</sup>/L) |  |  |
| Neutrophils | 4330 [3100;6390] * | 6344 |
| Lymphocytes | 950 [700;1300] * | 6371 |
| <800 | 2085 (32.7%) |  |
| 800-1000 | 987 (15.5%) |  |
| 1000-1200 | 1173 (18.4%) |  |
| >1200 | 2126 (33.4%) |  |
| Eosinophils | 0 [0;20] * | 6251 |
| Monocytes | 400 [300;600] * | 6271 |
| Hemoglobin (g/dL) | 13.9 [12.6;15] * | 6390 |
| Platelet count (x 10<sup>6</sup>/L) | 185000 [145000;240000] * | 6387 |
| **Arterial blood gases** |  |  |
| pH | 7.4 [7.4;7.5] * | 3400 |
| PCO2 (mmHg) | 34.2 [31;39] * | 3472 |
| PO2 (mmHg) | 66.9 [57;78.5] * | 3281 |
| PO2/FiO2 ratio (%) | 294.8 [241.1;347.6] * | 3185 |
| **Metabolic panel** |  |  |
| Glucose (mg/dL) | 110 [98;132] * | 6213 |
| Creatinine (mg/dL) | 0.9 [0.7;1.1] * | 6373 |
| Urea (mg/dL) | 36 [27;53] * | 5253 |
| Lactate dehydrogenase (U/L) | 304 [237;399] * | 5468 |
| <250 | 1646 (30.1%) |  |
| 250-400 | 2469 (45.2%) |  |
| >400 | 1353 (24.7%) |  |
| Aspartate aminotransferase (U/L) | 35 [25;52] * | 4903 |
| Alanine aminotransferase (U/L) | 29 [19;46] * | 5937 |
| C-reactive protein (mg/L) | 52 [15.7;115.5] * | 6093 |
| Lactate (mmol/L) | 1.5 [1.1;2.1] * | 3074 |
| Procalcitonin (ng/mL) | 0.1 [0.1;0.2] * | 3114 |
| Interleukin-6 (pg/mL) | 28 [10;59] * | 791 |
| D-dimer (ng/mL) |  | 4835 |
| <500 | 1863 (38.5%) |  |
| 500-1000 | 1540 (31.9%) |  |
| >1000 | 1432 (29.6%) |  |
| Serum ferritin (mcg/L) |  | 2135 |
| <300 | 589 (27.6%) |  |
| 300-650 | 591 (27.7%) |  |
| >650 | 955 (44.7%) |  |
| **qSOFA index** |  | 5931 |
| Low risk (≤1) | 5377 (90.7%) |  |
| High risk (≥2) | 554 (9.3%) |  |

Footnote: qSOFA: quick sequential organ failure assessment score.

Laboratory findings at admission are also shown in Table 2. Decreased lymphocytes and eosinophil counts were of note: the median values were 950 and 0 x 10^6^/L, respectively. High lactate dehydrogenase (LDH), D-dimer, and ferritin levels were observed in 12.4%, 61.5%, and 72.4%, respectively.

Treatment and complications during hospitalization are summarized in Table 3. A wide variety of drugs with purported antiviral effects have been used, the most frequent of which were hydroxychloroquine (85.7%) and lopinavir/ritonavir (62.4%). Remdesivir was only used in 28 patients (0.4%). Antibiotics were also widely indicated, mainly beta-lactam antibiotics (73.7%) and azithromycin (60.6%). Immunomodulatory drugs were also common, principally corticosteroids (32.9%), beta-interferon (13.3%), and tocilizumab (8.9%). Low-molecular-weight heparin was used in 80.6% of patients, generally at prophylactic doses. Many patients required respiratory support: high-flow nasal cannula was used in 7.9% of patients, noninvasive positive-pressure ventilation in 5.2%, and invasive mechanical ventilation in 5.6%. The main complication was acute respiratory distress syndrome (ARDS), which approximately one third of patients developed (32.3%), followed by bacterial pneumonia, and sepsis. Although 1,024 patients developed severe ARDS, only 483 (7.5%) were admitted to an intensive care unit.

**Table 3.** Treatment and complications during admission.

|  | Absolute frequency (%) | Valid cases |
| --- | --- | --- |
| **Antimicrobial therapy** |  |  |
| Hydroxychloroquine | 5467 (85.7%) | 6380 |
| Lopinavir/Ritonavir (LPV/r) | 3978 (62.4%) | 6372 |
| Azithromycin | 3845 (60.6%) | 6350 |
| Beta-lactam antibiotics | 4682 (73.7%) | 6353 |
| Remdesivir | 28 (0.4%) | 6294 |
| **Immunomodulatory therapy** |  |  |
| Systemic corticosteroids | 2083 (32.9%) | 6337 |
| Interferon Beta-1B (IFNβ) | 838 (13.3%) | 6320 |
| Tocilizumab | 564 (8.9%) | 6338 |
| Anakinra | 26 (0.4%) | 6209 |
| Immunoglobulin | 20 (0.3%) | 6027 |
| **Ventilation therapy** |  |  |
| High flow nasal cannula | 494 (7.9%) | 6290 |
| Invasive mechanic ventilation | 355 (5.6%) | 6353 |
| Non-invasive mechanical ventilation | 329 (5.2%) | 6354 |
| **Anticoagulant therapy** |  |  |
| Low-molecular-weight heparin therapy during hospitalization |  | 6318 |
| No | 1223 (19.4%) |  |
| Low (prophylactic) dose | 4082 (64.6%) |  |
| High (anticoagulant) dose | 623 (9.9%) |  |
| Intermediate dose | 390 (6.2%) |  |
| **Complications** |  |  |
| Acute respiratory distress syndrome |  | 6371 |
| No | 4305 (67.6%) |  |
| Mild | 568 (8.9%) |  |
| Moderate | 474 (7.4%) |  |
| Severe | 1024 (16.1%) |  |
| Bacterial pneumonia | 642 (10.1%) | 6382 |
| Sepsis | 382 (6.0%) | 6378 |
| Intensive care unit admission | 483 (7.5%) | 6411 |
| **Outcome** |  |  |
| Discharge | 5031 (78.3%) | 6424 |
| Death | 1356 (21.1%) | 6424 |
| Re-admission | 192 (3.3%) | 5742 |
| Not discharged at end of follow-up (after re-admission) | 37 (0.6%) | 6424 |

The median follow-up period was 40 days (range: 0-102 days). At the end of follow-up, 78.3% had been discharged, 21.2% had died, and 0.6% continued hospitalized (after re-admission). The average length of hospital stay for discharged patients was 10.4 days (range: 1-62 days). The rate of readmission within 30 days was 3.8% (192 patients out of 5,088 discharges).

## DISCUSSION

In this study, we analyze a large series of hospitalized patients with COVID-19 in Spain who have been included in the ongoing SEMI-COVID Network. This first cohort includes consecutive patients admitted to hospitals nationwide who were discharged or died.

Similar to almost all Western series, our patients were predominantly elderly, male, and with multiple comorbidities. Recently, the first conclusions about the impact of COVID-19 in Madrid, the epicenter of the pandemic in Spain, were drawn from a large cohort of 2,226 patients from La Paz University Hospital of Madrid by Borobia et al 2020 (unpublished data). Its strengths and weaknesses are both a result of its single-center design: the data are more consistent and able to be analyzed, but also less able to be extrapolated to the general population and prone to local biases, such as different population demographics or features specific to that particular hospital. Our series has a higher proportion of males, as has been described in most multicenter cohorts and contrary to the work by Borobia et al. The higher proportion of females at La Paz University Hospital may be a result of its specific demographic features and does not reflect the differences according to sex previously described in viral infections and specifically in COVID-19. In addition, our cohort comprises older patients with a greater number of comorbidities. In our series, the median age was 69 years (61 in Madrid cohort), which is clearly higher than Guan et al.’s Chinese series [4], moderately higher than Richardson et al.’s New York series [7], and lower than Docherty et al.’s UK series (data unpublished). The most frequent comorbidities (hypertension, diabetes, dementia, and others) are similar to those that have been previously described, but all were more prevalent among our patients (Table 4).

**Table 4.**
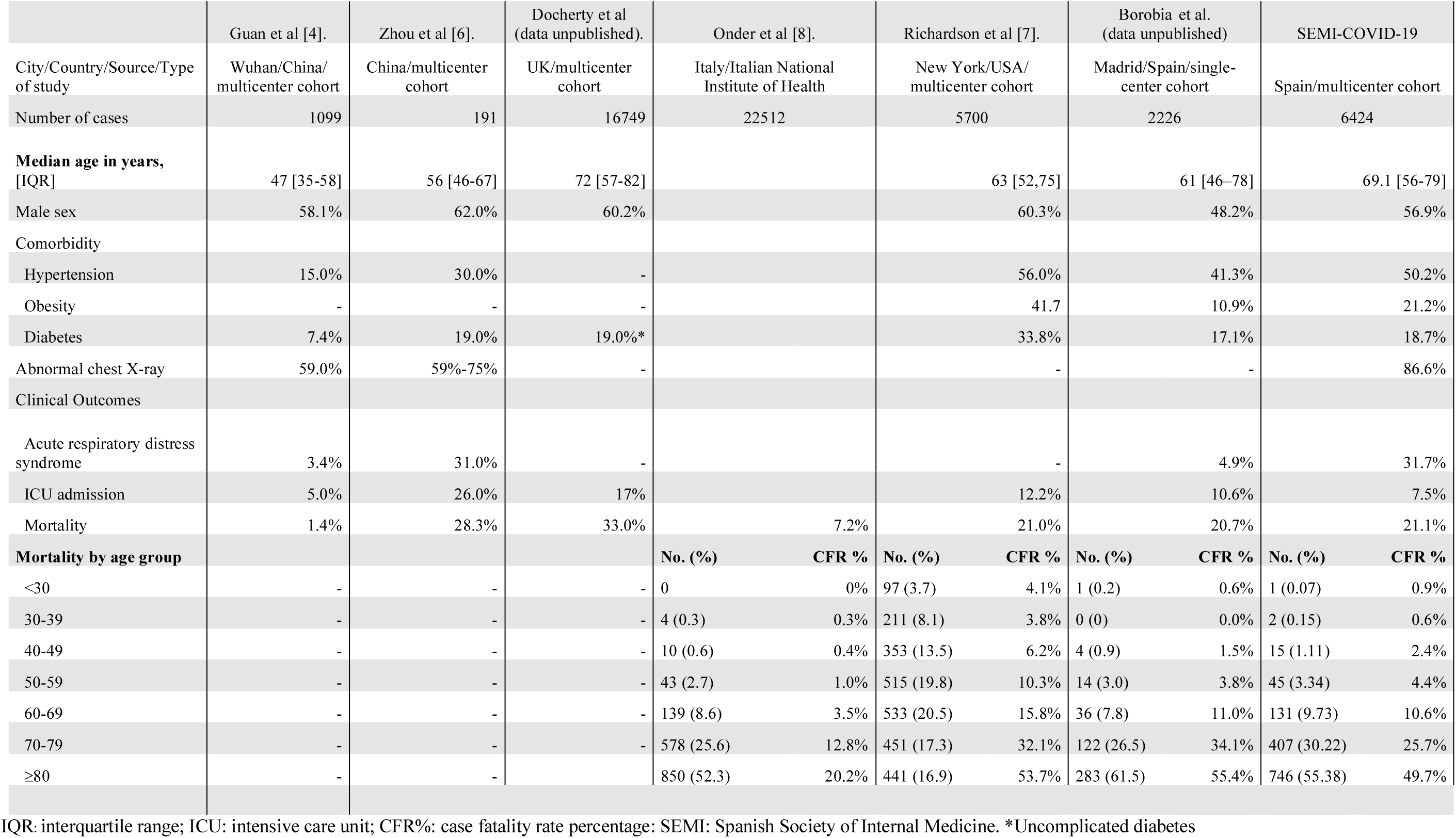
Comparison of baseline characteristics and outcome of patients with COVID-19 included in series from different countries.

In our cohort, symptoms reported upon arrival to the hospital (fever, cough, dyspnea, and asthenia) were similar to those reported in other studies [4-7], although myalgia and anosmia were less common. This could be explained by a potential difference in admission criteria, with smaller hospitals only admitting more severe cases and thus discharging patients without lung involvement from the emergency department.

In our series, mortality, as defined by CFR, was similar to what was observed in the Madrid cohort, some Chinese series [2-6], and the USA cohort [7], but was much higher than the one described in Italy [8] and lower than the figure observed in the UK. The difference in mortality rate between our series and the Italian series warrants some explanation, as we share many demographical features with Italy and the timing and magnitude of the COVID-19 pandemic have been similar.

It may reflect different selection criteria for the study or different hospital admission criteria. Less strict admission criteria yield a greater number of patients who meet the selection criteria, thus lowering the CFR. Indeed, population-based studies, which include more patients with less severe disease, have lower CFRs than hospital-centered series [8]. Conversely, stricter hospital admission criteria lead to fewer patients being included in the registry and increases CFR, as these patients have more severe disease. Additionally, factors related to race, including the percentage and origin of immigrant populations in each country, as well as healthcare-system disparities cannot be controlled for in these works. In fact, racial and demographic factors may be behind the differences in severity and outcomes between China and most Western countries[2-7].

Demographic factors, such as age or comorbidities, may partially explain the differences in mortality and can be controlled for by means of multivariate analysis. Pressure on the healthcare system can result in different mortality rates, as was shown in China by Liang et al. [11], who compared CFR both within and outside of Hubei province (CFR of 7.3% vs. 0.3%). In Italy [8], the highest disease burden was limited to the region of Lombardy whereas in Spain, it has been more widely distributed. Nevertheless, the majority of patients in our series are from Madrid, which has been one of the regions most affected by COVID-19 and where the situation is comparable to that of northern Italy. This will be further explored in ongoing studies.

As has been shown in all series, a high percentage of patients had abnormal laboratory values at admission that were consistent with an impaired immune-inflammatory profile [2-7]. In our series, lymphopenia and elevated levels of D-dimer, lactate dehydrogenase, and ferritin were the most frequent findings. Also, most patients received treatment that is purportedly effective against SARS-CoV-2. Our multicenter registry has been designed to allow for multivariate analysis of the prognostic value of these abnormal laboratory findings as well as treatment received during hospitalization.

Notably, in our series, there was a much higher proportion of patients with ARDS (moderate or severe: 23.5%) than patients who were admitted to an ICU (7.5%). This suggest that only one out of every three patients with ARDS was admitted to an ICU. We have discussed this finding in detail and have evaluated some obvious confounding factors and biases. Patients admitted directly to an ICU or who died in an ICU could have been lost to our cohort and thus altered our ICU admission ratio. Patients still in an ICU who have not been discharged or died at the end of follow-up on this cohort are not included in our registry and thus also falsely lower our ICU admission rate. But it still does not explain how 541 out of 1024 patients with ARDS were not admitted to ICU, not all of them dying. Another plausible explanation is healthcare system overload, at least in the most affected parts of the country. For instance, the number of ICU beds has increased substantially during the COVID-19 pandemic in Spain [1]. It is likely that in addition to increasing ICU capacity, some semi-intensive care areas were established within hospitals in order to provide intensive care in wards outside of the ICU. In our personal experience, most hospitals have designed specialized out-of-ICU semi-intensive or intermediate care wards in order to provide respiratory support to patients when ICU expansion was no longer feasible. This finding warrants further examination.

Our collaborative effort has provided us with a large amount of data from a sizeable number of patients. Among the strengths of this study are its multicenter design; its wide geographical dispersion, which limits local biases (selection, admission, treatment availability, ICU availability); and its large size, which provides statistical power for confirming hypotheses.

This study also has a number of limitations. First, data are collected by a large team of researchers, which could lead to heterogeneity in data input and validation. Second, the registry includes consecutive patients from participating centers, which limits patient selection bias but introduces another selection bias according to participating centers. Third, our registry, though extensive (more than 300 variables), provides only basic data for enhancing our knowledge of COVID-19, but lacks the level of detail required for deeper analysis. Lastly, the main limitation of this study is its observational design, which does not allow for establishing causal inferences.

This is the largest reported series of hospitalized patients in Spain with confirmed COVID-19 infection and one of the largest registries in the world to date. Though our findings are currently preliminary and must be explored in further detail and greater depth, the ongoing SEMI-COVID Network will surely become a key tool to help clinicians and scientists improve knowledge of this novel disease which has threatened not only the lives of many patients and the operations of our healthcare systems, but also the foundations of our economy and way of life.

## Data Availability

NA

## ACKNOWLEDGEMENTS

We gratefully acknowledge all the investigators who participate in the SEMI-COVID-19 Network. We also thank the SEMI-COVID-19 Registry Coordinating Center, S&H Medical Science Service, for their quality control data, logistic and administrative support. The authors declare that there are no conflicts of interest.

## REFERENCES

1. España: Ministerio de Sanidad. Situación de COVID-19 en España. [Internet]. Centro de Coordinación de Alertas y Emergencias Sanitarias. Enfermedad por el coronavirus (COVID-19). 2020 [cited 2020 May 8]. Available from: https://covid19.isciii.es/

2. Wang D, Hu B, Hu C, Zhu F, Liu X, Zhang J, et al. Clinical Characteristics of 138 Hospitalized Patients with 2019 Novel Coronavirus-Infected Pneumonia in Wuhan, China. JAMA. 2020;323(11):1061–1069. doi:10.1001/jama.2020.1585

3. Chen N, Zhou M, Dong X, Qu J, Gong F, Han Y, et al. Epidemiological and clinical characteristics of 99 cases of 2019 novel coronavirus pneumonia in Wuhan, China: a descriptive study. Lancet. 2020;395(10223):507–513. doi:10.1016/S0140-6736(20)30211-7.

4. Guan W, Ni Z, Hu Y, Liang W, Ou C, He J, et al. Clinical Characteristics of Coronavirus Disease 2019 in China. N Engl J Med. 2020;382(18):1708–1720. doi:10.1056/NEJMoa2002032

5. Huang C, Wang Y, Li X, Ren L, Zhao J, Hu Y, et al. Clinical features of patients infected with 2019 novel coronavirus in Wuhan, China. Lancet. 2020;395(10223):497–506. doi:10.1016/S0140-6736(20)30183-5

6. Zhou F, Yu T, Du R, Fan G, Liu Y, Liu Z, et al. Clinical course and risk factors for mortality of adult inpatients with COVID-19 in Wuhan, China: a retrospective cohort study. Lancet. 2020;395(10229):1054–1062. doi:10.1016/S0140-6736(20)30566-3

7. Richardson S, Hirsch JS, Narasimhan M, Crawford JM, McGinn T, Davidson KW, et al. Presenting Characteristics, Comorbidities, and Outcomes Among 5700 Patients Hospitalized With COVID-19 in the New York City Area. JAMA. 2020;e206775. doi:10.1001/jama.2020.6775

8. Onder G, Rezza G, Brusaferro S. Case-Fatality Rate and Characteristics of Patients Dying in Relation to COVID-19 in Italy JAMA. 2020; 10.1001/jama.2020.4683. doi:10.1001/jama.2020.4683

9. NIH Interim Guidelines Coronavirus Disease COVID-19 [Internet]. [cited 2020 Apr 28]. Available from: https://www.covid19treatmentguidelines.nih.gov/introduction/

10. Liang WH, Guan WJ, Li CC, Li YM, Liang HR, Zhao Y, et al. Clinical characteristics and outcomes of hospitalised patients with COVID-19 treated in Hubei (epicenter) and outside Hubei (non-epicenter): A Nationwide Analysis of China. Eur Respir J. 2020 Apr 8:2000562. doi: 10.1183/13993003.00562-2020

